# Multifocal transient cortical brain lesions: a consistent MRI finding in neuro-COVID-19 patients

**DOI:** 10.1101/2020.05.19.20103168

**Authors:** Nicoletta Anzalone, Antonella Castellano, Roberta Scotti, Anna Mara Scandroglio, Massimo Filippi, Fabio Ciceri, Moreno Tresoldi, Andrea Falini

**Author notes:** Corresponding Author: Nicoletta Anzalone, MD Department of Neuroradiology IRCCS San Raffaele Scientific Institute and Vita-Salute San Raffaele University Via Olgettina 60 20132 Milan, Italy.

## Abstract

We report four cases of subacute encephalopathy occurring in patients with SARS-CoV-2 infection. All patients have been intubated in the first week from onset of ARDS and presented neurological signs of agitation and spatial disorientation after weaning from mechanical ventilation. The MRI picture and the clinical features are described. MRI lesions characteristics are unusual but demonstrate a highly consistent pattern through all the four patients with similar neurological symptoms. They do not fulfill any typical criteria for a definite neuroradiological entity. Their predominantly parieto-occipital distribution recalls posterior reversible encephalopathy syndrome (PRES), although the prevalent cortical involvement and diffusion MRI pattern are not typical of PRES. We speculate that this pattern may be related to a possible transient dysregulation of vasomotor reactivity.

---

Dear Sirs,

Neurologic manifestations of severe acute respiratory syndrome coronavirus 2 (SARS-CoV-2) have been recently reported [1,2], with relevance to vascular aetiology [2,3]. The neuroinvasive potential of SARSCoV-2 has also been advocated [4], possibly supporting isolated findings of encephalitis in COVID-19 patients [5]. More recently, acute necrotizing encephalopathy (ANE) related to intracranial cytokine storms has been reported, causing blood-brain-barrier breakdown without direct viral invasion [6].

Here we report four cases of subacute encephalopathy occurring in patients with SARS-CoV-2 infection. They are part of a series of 21 patients presenting with neurological symptoms studied with brain MRI with otherwise no significant imaging findings.

A multifocal involvement of the cortex was evident in all cases (Fig. 1 and 2). The multiple areas, from punctiform to some millimeters in extension, appeared hyperintense on T2-weighted and FLAIR images and were located in the parietal, occipital and frontal regions. On diffusion MRI, all but two of the lesions were characterized by the absence of apparent diffusion coefficient (ADC) changes (Fig. 1c and Fig. 2c). A minimum involvement of the adjacent subcortical white matter was evident in only a few lesions. Very subtle contrast enhancement was detected only in a cortical lesion. In one patient a follow-up MRI scan was obtained after one month, demonstrating a complete resolution of all the lesions (Fig. 2f-l).

**Fig 1.**
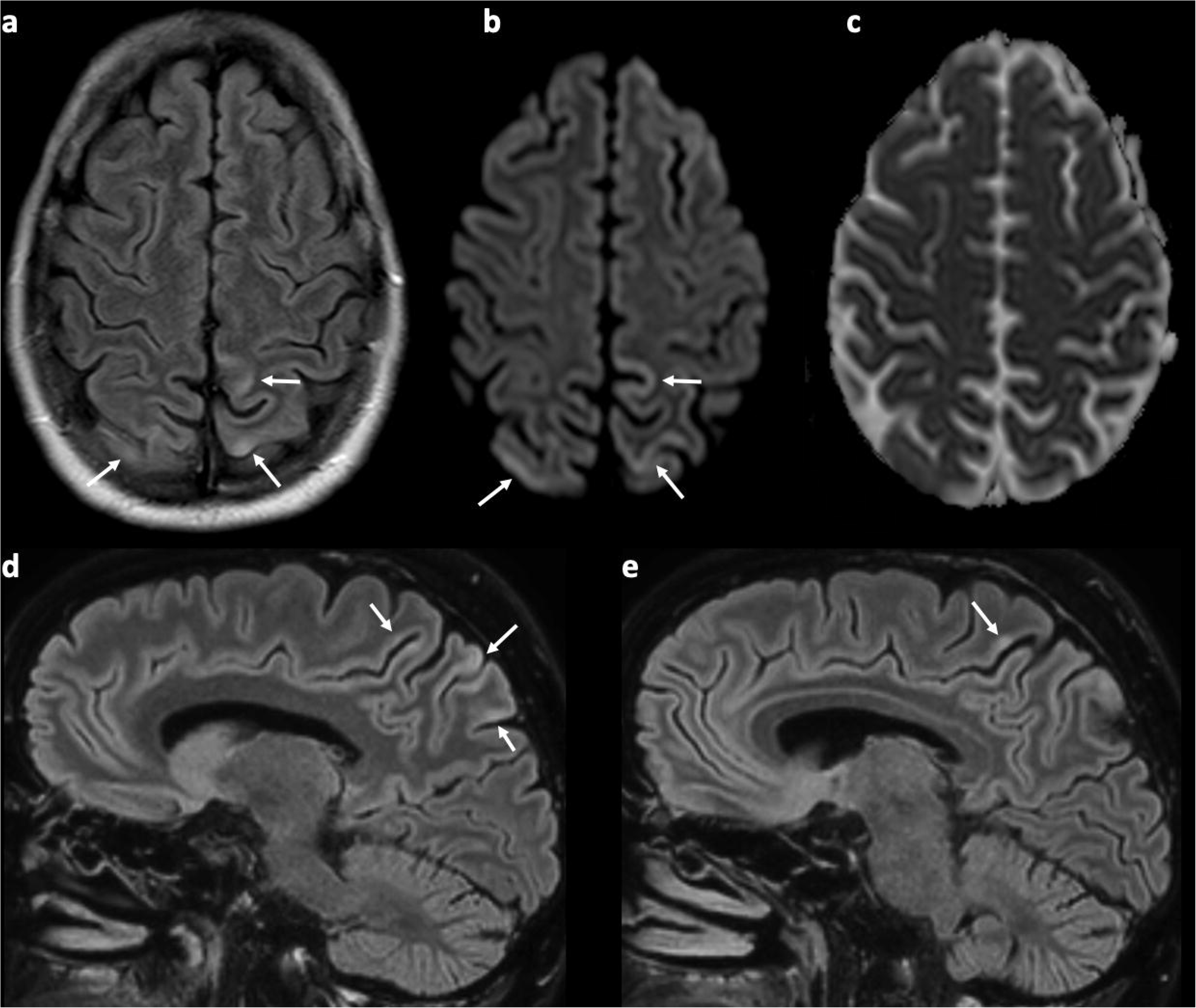
47-years-old man diagnosed with COVID-19 and presenting neurological signs of agitation and spatial disorientation after weaning from mechanical ventilation. Before the onset of neurological symptoms, laboratory findings revealed a C-reactive protein peak (262.5 mg/L, normal range, 0–6 mg/L). (a) Axial FLAIR, (b) diffusion-weighted image (DWI), (c) apparent diffusion coefficient (ADC) map and (d, e) sagittal FLAIR MR images. Multiple, cortical areas of punctiform and gyriform FLAIR and DWI hyperintensity (arrows) in both parietal lobes, with no ADC changes.

**Fig 2.**
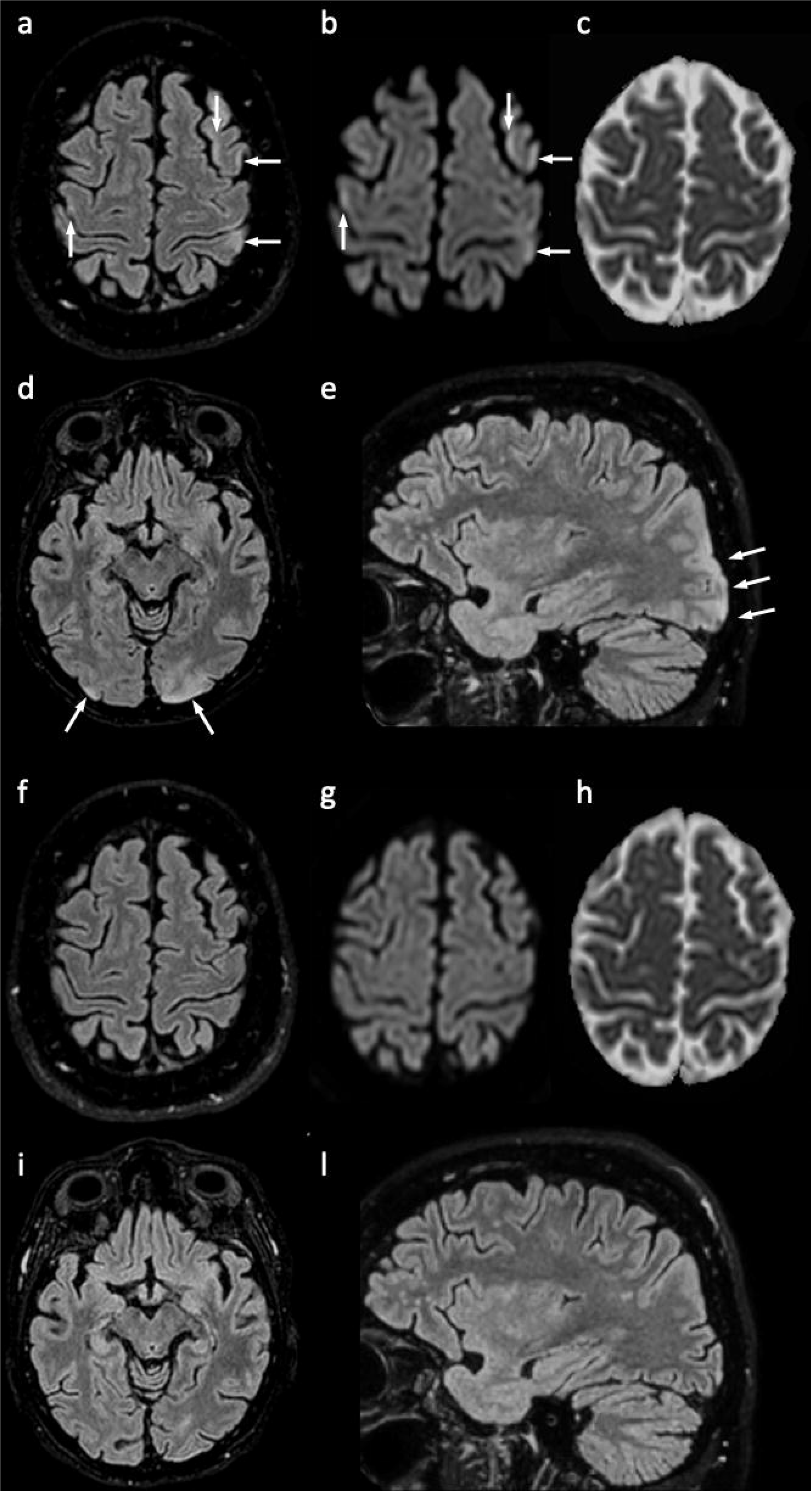
54-years-old woman diagnosed with COVID-19 and presenting neurological signs of agitation and spatial disorientation after weaning from mechanical ventilation. Before the onset of neurological symptoms, laboratory findings revealed a small C-reactive protein peak (27.5 mg/L, normal range, 0–6 mg/L) and raise of total white blood cell count (12.9 × 10^9^/L, normal range, 4.0–10.0 × 10^9^/L). Cerebrospinal fluid (CSF) analysis performed on the same day was negative for the presence of SARS-CoV-2 viral nucleic acid. (a-e) Initial MRI scan. (f-l) Follow-up MRI after one month. (a, d, f, i) Axial FLAIR, (b, g) diffusion-weighted image (DWI), (c, h) apparent diffusion coefficient (ADC) map and (e, l) sagittal FLAIR MR images. Multifocal linear and punctiform cortical FLAIR and DWI hyperintensities in the left parietal lobe, bilateral precentral gyri and left middle frontal gyrus (a-c), with no ADC changes. Bilateral occipital involvement is shown in (d, e), with a cortical/subcortical FLAIR hyperintense lesion at the level of the left occipital pole. (f-l) Follow-up MRI demonstrates a complete resolution of all the lesions.

All patients (2 men, 2 women; age range: 46–63 years) have been intubated in the first week from onset of ARDS and presented neurological signs of agitation and spatial disorientation after weaning from mechanical ventilation. One patient had a generalized seizure. The time interval from onset of neurological symptoms to MRI was 2–6 days. Diagnosis of COVID-19 was made by detection of SARS-CoV-2 viral nucleic acid in a nasopharyngeal swab specimen. All patients received the same treatment for SARS-CoV-2 infection. None of the patients had a relevant clinical history or previous treatment or hypertension. Laboratory findings revealed in all cases a second smaller C-reactive protein peak from the initial one and raise of serum level of aspartate and alanine transaminase before the onset of neurological symptoms. D-dimer elevation was present and stable during the disease course.

MRI lesions’ characteristics are unusual but demonstrate a highly consistent pattern through all the four patients with similar neurological symptoms. They do not fulfill any typical criteria for a definite neuroradiological entity. Although their predominantly parieto-occipital distribution recalls posterior reversible encephalopathy syndrome (PRES) [7], the prevalent cortical involvement and diffusion MRI pattern are not typical of PRES. We speculate that this pattern may be related to a possible transient dysregulation of vasomotor reactivity. In particular, the cortical involvement may suggest a possible vascular mechanism more shifted toward transient vasoconstriction.

It is currently known that SARS-CoV-2 might dysregulate the renin-angiotensin system (RAS) system by acting on ACE2 receptors, causing microcirculation impairment possibly impacting on blood flow regulation. More recently, evidence of direct viral infection of the endothelial cell and diffuse endothelial inflammation has been reported, resulting in endothelial dysfunction and impaired microcirculatory function [8]. Along with inflammation, there is a tendency to thrombosis in more severe cases [9]. Nonetheless, other vasculo-mediated mechanisms including altered vasomotor reactivity may play a role and cause neurological symptoms in COVID-19 patients [8]. In this regard, normalization of MRI findings in one patient (Figure 2f-1) may corroborate the hypothesis of a transient functional nature of the impaired cerebral microcirculatory function.

We believe that, due to the peculiarity and subtle appearance of the MRI findings, our report may alert neurologists and radiologists to the existence of this subacute neuroimaging picture in SARS-CoV-2 patients, clearly different from cortical ischemia, encephalitis or acute necrotizing encephalopathy, and also to inform clinicians about the possible spontaneous reversibility of the picture.

## Data Availability

Data are available upon request.

## Author contributions

NA, AC, RS, AF: acquisition of data, analysis and interpretation of data, drafted and revised the manuscript for intellectual content. AMS, MF, FC, MT: interpretation of data, revised the manuscript for intellectual content.

## Compliance with ethical standards

### Ethical approval

The study was approved by the Ethics Committee of IRCCS San Raffaele Scientific Institute, Milan, Italy.

### Informed consent

All patients provided signed informed consent prior to MR imaging. Informed consent was collected from the patients for the inclusion of deidentified clinical data in a scientific publication, in accordance with the Declaration of Helsinki.

